# Biomarker panels for improved risk prediction and enhanced biological insights in patients with atrial fibrillation

**DOI:** 10.1101/2024.12.19.24319346

**Authors:** Pascal B. Meyre, Stefanie Aeschbacher, Steffen Blum, Tobias Reichlin, Moa Haller, Nicolas Rodondi, Andreas S. Müller, Alain Bernheim, Jürg Hans Beer, Giorgio Moschovitis, André Ziegler, Bianca Wahrenberger, Elia Rigamonti, Giulio Conte, Philipp Krisai, Leo H. Bonati, Stefan Osswald, Michael Kühne, David Conen

**Affiliations:** Division of Cardiology, Department of Medicine, University Hospital Basel, Switzerland; Cardiovascular Research Institute Basel, University Hospital Basel, Switzerland; Department of Cardiology, Inselspital, Bern University Hospital, 3010 Bern, Switzerland; Institute of Primary Health Care (BIHAM), University of Bern, Bern, Switzerland; Department of General Internal Medicine, Inselspital, Bern University Hospital, University of Bern, Bern, Switzerland; Department of Cardiology, Triemli Hospital Zurich, Zurich, Switzerland; Department Internal Medicine, Baden Switzerland and Center of Molecular Cardiology, Cantonal Hospital Baden, University of Zürich, Zürich, Switzerland; Divison of Cardiology, Regional Hospital of Lugano, Ente Ospedaliero Cantonale (EOS), Lugano, Switzerland; Cardiocentro Ticino Institute, Ente Ospedaliero Cantonale (EOC), Lugano, Switzerland; Roche Diagnostics International, Rotkreuz, Switzerland; Rheinfelden Rehabilitation Clinic, Rheinfelden, Switzerland; Population Health Research Institute, McMaster University, Hamilton, ON, Canada; Department of Medicine, McMaster University, Hamilton, ON, Canada; Department of Health Research Methods, Evidence, and Impact, McMaster University, Hamilton, ON, Canada

**Author notes:** Corresponding author: Pascal B. Meyre, MD, PhD, Department of Cardiology, University Hospital of Basel, Cardiovascular Research Institute Basel, Petersgraben 4, CH-4031 Basel, Switzerland E.

## Abstract

Atrial fibrillation (AF) is associated with an increased risk of adverse cardiovascular events, but the underlying biological mechanisms remain incompletely understood. Here we evaluated a panel of 12 circulating biomarkers representing diverse pathophysiological pathways in a cohort of 3,817 AF patients to assess their association with adverse cardiovascular outcomes. We identified 5 biomarkers—d-dimer, growth differentiation factor 15 (GDF-15), interleukin-6 (IL-6), N-terminal pro-B-type natriuretic peptide (NT-proBNP), and high-sensitivity troponin T (hsTropT)—that were independently associated with cardiovascular death, stroke, myocardial infarction, and systemic embolism, significantly enhancing predictive accuracy. Additionally, GDF-15, insulin-like growth factor-binding protein-7 (IGFBP-7), NT-proBNP, and hsTropT were strong predictors of heart failure hospitalization, while GDF-15 and IL-6 were associated with major bleeding events. Incorporating IL-6, NT-proBNP, and hsTropT to the CHA₂DS₂-VASc score improved stroke risk prediction. Machine learning models incorporating these biomarkers demonstrated consistent improvements in risk stratification across all outcomes. Our results highlight the potential of integrating biomarkers related to myocardial injury, inflammation, oxidative stress, and coagulation into both conventional and machine learning-based models refine prognosis and guide clinical decision-making in AF patients.

---

Patients with atrial fibrillation (AF) have an increased risk of death, cardiovascular events, and bleeding. Findings from prospective cohort studies suggest that AF is associated with an approximately 5-fold increased risk of stroke, 2-fold increased risk of myocardial infarction and death ^1–3^, and at least a 3-fold increased risk of heart failure ^1,4^. Additionally, AF patients have an increased risk of major bleeding due to oral anticoagulation treatment ^5^. As these complications carry significant risks for the individual patient and induce a significant burden to the healthcare system, it is of substantial clinical relevance to gain a better understanding for their underlying pathophysiology and to improve risk prediction. Recent histological analyses from AF biopsies showed that the atrial tissue undergoes extensive changes over time, due to inflammation, blood clotting, vascular permeability and collagen infiltration ^6^. Circulating biomarkers that reflect these pathological pathways offer a promising, less invasive approach to assessing their involvement in individual patients. The aim of this study was to identify biomarker patterns using a panel of selected biomarkers that reflect distinct disease pathways associated with major adverse cardiac events and bleeding in AF patients. By leveraging both traditional statistical approaches and machine learning models, we sought to deepen our understanding of AF pathophysiology and enhance risk prediction for major cardiovascular events and bleeding complications in this population.

## Results

A total of 3,817 AF patients were included in this analysis. Mean age was 71 ± 10 years and 28% were female **(Table 1)**. Nearly half had paroxysmal AF (49%), and non-paroxysmal forms of AF were present in 51% (28% persistent and 23% permanent AF). Hypertension was the most common cardiovascular risk factor (69%), followed by coronary artery disease (27%) and heart failure (24%). Overall, 84% of patients were on oral anticoagulation. Spearman rank correlations between biomarkers are presented in **Supplementary Figure 3**. hsTropT showed strong correlations (>0.60) with GDF-15 (0.64) and cystatin C (0.62). OPN was strongly correlated with cystatin C (0.77) and GDF-15 (0.64). IGFBP-7 exhibited strong correlations with cystatin C (0.68) and GDF-15 (0.67). NT-proBNP had a strong correlation with ANG-2 (0.69).

**Table 1.** Baseline characteristics.

| Characteristic | N=3817 |
| --- | --- |
| Age, years | 71±10 |
| Female sex | 1067 (28) |
| Body mass index, kg/m<sup>2</sup> | 27±5 |
| Systolic blood pressure, mmHg | 134±19 |
| Heart rate, bpm | 70±17 |
| CHA<sub>2</sub>DS<sub>2</sub>-VASc score | 3.2±1.7 |
| Smoking status |  |
| Active | 298 (8) |
| Past | 1836 (48) |
| Never | 1673 (44) |
| Type of atrial fibrillation |  |
| Paroxysmal | 1869 (49) |
| Persistent | 1070 (28) |
| Permanent | 875 (23) |
| Medical history |  |
| Hypertension | 2635 (69) |
| Diabetes | 614 (16) |
| Prior stroke/TIA | 651 (17) |
| Coronary artery disease | 1021 (27) |
| Prior myocardial infarction | 564 (15) |
| Peripheral vascular disease | 281 (7) |
| Heart failure | 909 (24) |
| Major bleeding | 146 (4) |
| Chronic kidney disease | 708 (19) |
| Oral anticoagulation | 3212 (84) |
| Direct oral anticoagulants | 1346 (35) |
| Vitamin K antagonists | 1864 (49) |
| Data are presented as means ± standard deviations or counts (percentages). Abbreviations: TIA, transient ischemic attack. |  |

To identify biomarkers associated with cardiovascular events, we conducted age- and sex-adjusted, and multivariable-adjusted Cox regression analyses for each biomarker and outcome **(Supplementary Tables 5-14 and in Supplementary Figure 4)**. For the composite outcome, 5 biomarkers including d-dimer, GDF-15, IL-6, NT-proBNP and hsTropT independently contributed to the model fit **(Supplementary Table 5, Figure 1A)**. Notably, hsTropT, NT-proBNP, and GDF-15 were among the most significant variables in the model **(Figure 2A)**. For heart failure hospitalization, 4 biomarkers - GDF-15, IGFBP-7, NT-proBNP and hsTropT - were significantly associated with the outcome **(Supplementary Table 6, Figure 1B)**, with NT-proBNP and GDF-15 being among the most important risk predictors **(Figure 2B)**. GDF-15, IGFBP-7, IL-6 and hsTropT were associated with an increased risk of major bleeding events **(Supplementary Table 7, Figure 1C)**, with GDF-15 being one of the most important biomarkers in the model **(Figure 2C)**. NT-proBNP and IL-6 were associated with both ischemic and the composite of ischemic and hemorrhagic stroke **(Supplementary Tables 8 and 9, Figures 1D** and **1E)**, and NT-proBNP was among the most important risk indicators **(Figure 2D** and **2E)**. Both IL-6 and hsTropT were linked to a higher risk of MI **(Supplementary Table 10, Figure 1F)**, and both were among the major predictors in the model **(Figure 2F)**. For cardiovascular death, GDF-15, IL-6, NT-proBNP and hsTropT were associated with the outcome **(Supplementary Table 11, Figure 1G)**. GDF-15, NT-proBNP and hsTropT were the most important predictors **(Figure 2G)**. D-dimer, GDF-15, IGFBP-7, IL-6, NT-proBNP and hsTropT were associated with all-cause death **(Supplementary Table 12, Figure 1H)**. GDF-15, IL-6 and hsTropT were key risk predictors **(Figure 2H)**. For the composite bleeding outcome and clinically relevant NM bleeding, GDF-15, IL-6 for composite bleeding and NT-proBNP for clinically relevant NM bleeding were associated with the outcomes **(Supplementary Tables 13 and 14**, **Figure 1I** and **1J)**. GDF-15 was the most important risk indicator in the model **(Figure 2I** and **2J)**.

**Figure 1.**
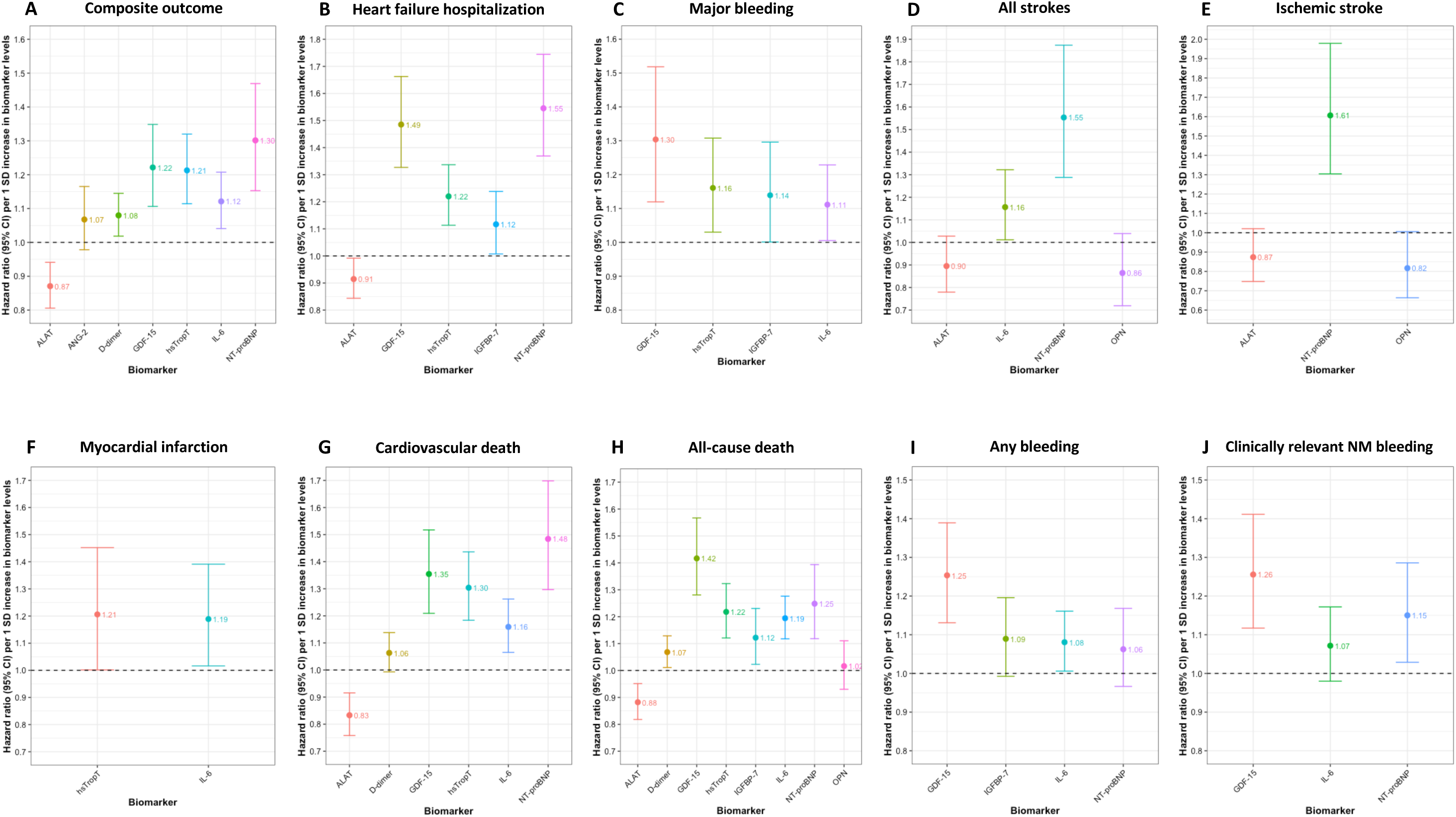
Associations between selected biomarkers and adverse cardiac outcomes from combined Cox models. This figure shows standardized hazard ratios and 95% CIs of associations between backward-selected biomarkers and different adverse cardiac outcomes, derived from combined multivariable Cox models.

**Figure 2.**
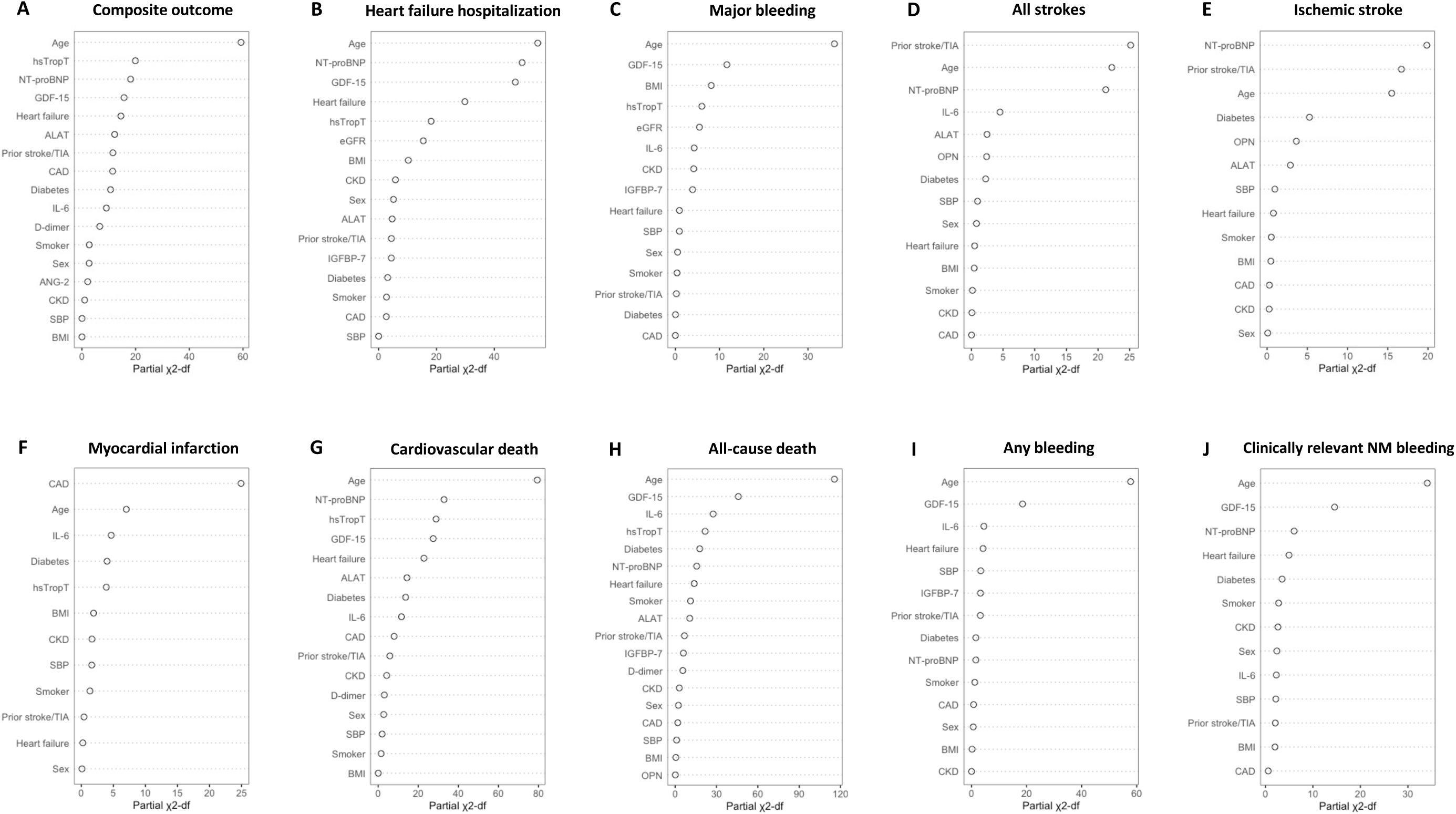
Relative importance of predictors from combined Cox models. This figure shows the relative importance of each clinical variable and backward-selected biomarkers for different adverse cardiac outcomes, derived from combined multivariable Cox models. Abbreviations: ALAT=Alanine aminotransferase; ANG-2=Angiopoetin-2; BMI=body mass index; CAD=coronary artery disease; CKD=chronic kidney disease; eGFR=estimated glomerular filtration rate; GDF-15=growth differentiation factor-15; hsTropT=high-sensitivity troponin T; IGFBP-7= Insulin-like growth factor-binding protein-7; IL-6= Interleukin-6; NT-proBNP= N-terminal pro-B-type natriuretic peptide; OPN=Osteopontin; SBP=Systolic blood pressure.

We assessed the discriminatory performance of CHA_2_DS_2_-VASc score, both alone and in combination with biomarkers, for predicting stroke outcomes **(Figure 3)**. Following backward selection, IL-6, NT-proBNP and hsTropT were identified as independent predictors to model fit. For the composite of all strokes, the addition of biomarkers (IL-6, NT-proBNP and hsTropT) significantly improved risk prediction, with an increase in AUC from 0.64 to 0.66 (P=0.04 n=241 events). In contrast, the inclusion of NT-proBNP did not significantly improve risk prediction for ischemic stroke (AUC 0.63 vs 0.65, P=0.10, n=192 events).

**Figure 3.**
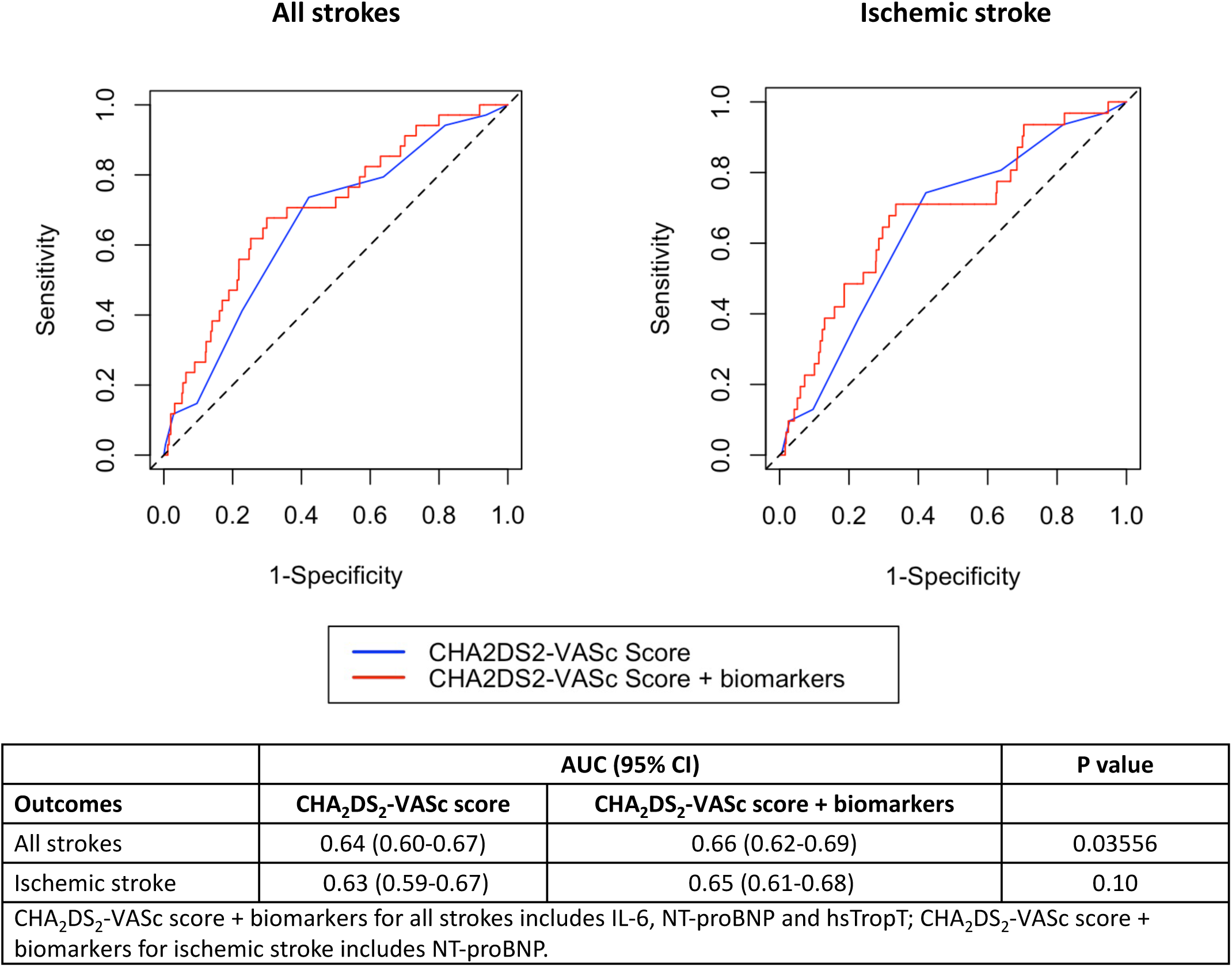
Discriminatory ability of CHA_2_DS_2_-VASc score with and without biomarkers for all strokes and ischemic stroke. This figure shows ROC curves of the CHA_2_DS_2_-VASc score with and without biomarkers for all strokes and ischemic stroke. Abbreviations: IL-6= Interleukin-6; NT-proBNP= N-terminal pro-B-type natriuretic peptide; hsTropT=high-sensitivity troponin T.

We investigated the impact of adding biomarkers to traditional Cox models and machine learning algorithms on cardiovascular risk prediction **(Table 2, Supplementary Figure 5)**. For composite outcome, the inclusion of biomarkers significantly improved model performance, increasing the AUC of the combined Cox model from 0.74 to 0.77 (P=2.6x10^-8^). The Random Forest model showed an increase in AUC from 0.74 to 0.75 (P=0.03), while the XGBoost model demonstrated an improvement from 0.95 to 0.97 (P=0.0007345). For heart failure hospitalization, the inclusion of biomarkers enhanced predictive accuracy across all models. The combined Cox model’s AUC increased from 0.77 to 0.80 (P=5.5x10^-10^), the LASSO model from 0.80 to 0.83 (P=0.04), the Random Forest model from 0.77 to 0.80 (P=0.0002564), and the XGBoost model from 0.96 to 0.98 (P=5.0x10^-6^). For major bleeding events, the addition of biomarkers resulted in improvements in some models. The combined Cox model’s AUC increased from 0.67 to 0.68 (P=0.01), and the Random Forest model showed a small but non-significant increase from 0.63 to 0.65 (P=0.10). However, the LASSO model did not show a significant change (AUC 0.69 to 0.70, P=0.50). In contrast, the XGBoost model exhibited an improvement, with the AUC increasing from 0.94 to 0.97 (P=8.8x10^-5^). Similar trends were observed for all secondary outcomes.

**Table 2.** Discriminative ability of Cox and machine-learning models for outcomes with and without biomarkers.

| **Outcomes** | **Model** | **AUC<sub>Base</sub> (95% CI)** | **AUC<sub>Base+biomarkers</sub> (95% CI)** | **P value** |
| --- | --- | --- | --- | --- |
| Composite | Combined Cox model | 0.74 (0.72-0.76) | 0.77 (0.75-0.79) | 2.6x10<sup>-8</sup> |
|  | LASSO | 0.77 (0.75-0.79) | 0.78 (0.76-0.80) | 0.3235 |
|  | Random forest | 0.74 (0.72-0.76) | 0.75 (0.73-0.78) | 0.02796 |
|  | XGBoost | 0.95 (0.94-0.96) | 0.97 (0.96-0.96) | 0.0007345 |
| HF hospitalization | Combined Cox model | 0.77 (0.75-0.78) | 0.80 (0.79-0.82) | 5.5x10<sup>-10</sup> |
|  | LASSO | 0.80 (0.78-0.82) | 0.83 (0.81-0.84) | 0.04671 |
|  | Random forest | 0.77 (0.74-0.79) | 0.80 (0.78-0.82) | 0.0002564 |
|  | XGBoost | 0.96 (0.95-0.97) | 0.98 (0.97-0.98) | 5.0x10<sup>-6</sup> |
| Major bleeding | Combined Cox model | 0.67 (0.64-0.69) | 0.68 (0.66-0.71) | 0.01352 |
|  | LASSO | 0.69 (0.66-0.72) | 0.70 (0.67-0.73) | 0.5016 |
|  | Random forest | 0.63 (0.60-0.66) | 0.65 (0.62-0.68) | 0.1015 |
|  | XGBoost | 0.94 (0.93-0.96) | 0.97 (0.96-0.98) | 8.8x10<sup>-5</sup> |
| Ischemic stroke | Combined Cox model | 0.65 (0.61-0.69) | 0.67 (0.63-0.71) | 0.03113 |
|  | LASSO | 0.68 (0.63-0.72) | 0.69 (0.64-0.73) | 0.7793 |
|  | Random forest | 0.59 (0.54-0.64) | 0.59 (0.54-0.64) | 0.8381 |
|  | XGBoost | 0.95 (0.93-0.97) | 0.98 (0.97-0.99) | 5.6x10<sup>-5</sup> |
| Any stroke | Combined Cox model | 0.66 (0.63-0.70) | 0.68 (0.65-0.72) | 0.02533 |
|  | LASSO | 0.69 (0.65-0.73) | 0.70 (0.66-0.74) | 0.8273 |
|  | Random forest | 0.61 (0.57-0.66) | 0.61 (0.57-0.65) | 0.8254 |
|  | XGBoost | 0.95 (0.94-0.96) | 0.97 (0.96-0.98) | 0.01199 |
| MI | Combined Cox model | 0.70 (0.66-0.74) | 0.70 (0.66-0.74) | 0.943 |
|  | LASSO | 0.75 (0.71-0.80) | 0.74 (0.70-0.79) | 0.7338 |
|  | Random forest | 0.68 (0.63-0.72) | 0.62 (0.57-0.67) | 0.0009897 |
|  | XGBoost | 0.97 (0.96-0.99) | 0.99 (0.98-0.99) | 0.02063 |
| CV death | Combined Cox model | 0.79 (0.77-0.81) | 0.83 (0.81-0.85) | $5.6 \times 10^{-14}$ |
|  | LASSO | 0.82 (0.80-0.85) | 0.85 (0.83-0.87) | 0.09099 |
|  | Random forest | 0.79 (0.77-0.82) | 0.82 (0.80-0.85) | 0.0001046 |
|  | XGBoost | 0.97 (0.96-0.98) | 0.98 (0.98-0.99) | 0.0004972 |
| All-cause death | Combined Cox model | 0.80 (0.78-0.81) | 0.84 (0.83-0.86) | $<2.2 \times 10^{-16}$ |
|  | LASSO | 0.82 (0.80-0.84) | 0.85 (0.83-0.87) | 0.01385 |
| | Random forest | 0.80 (0.78-0.82) | 0.83 (0.81-0.85) | $1.9 \times 10^{-7}$ |
| | XGBoost | 0.95 (0.94-0.96) | 0.98 (0.97-0.98) | $5.6 \times 10^{-10}$ |
| Any bleeding | Combined Cox model | 0.64 (0.62-0.66) | 0.65 (0.63-0.67) | 0.07765 |
|  | LASSO | 0.66 (0.64-0.69) | 0.68 (0.66-0.71) | 0.189 |
|  | Random forest | 0.63 (0.61-0.66) | 0.64 (0.62-0.67) | 0.2455 |
|  | XGBoost | 0.92 (0.91-0.94) | 0.92 (0.91-0.94) | 0.8432 |
| CRNMB | Combined Cox model | 0.61 (0.59-0.64) | 0.62 (0.60-0.64) | 0.2217 |
|  | LASSO | 0.66 (0.63-0.68) | 0.66 (0.63-0.69) | 0.8349 |
|  | Random forest | 0.60 (0.58-0.63) | 0.63 (0.60-0.65) | 0.05211 |
|  | XGBoost | 0.94 (0.93-0.96) | 0.96 (0.95-0.97) | 0.007685 |
| Cox base model includes age, sex, body mass index, current smoker, systolic blood pressure, history of diabetes, prior stroke or TIA, history of heart failure, chronic kidney disease and coronary artery disease.<br>Base models from machine learning models (LASSO, Random forest, XGBoost) includes all variables listed in the Supplementary Table 2. |  |  |  |  |

## Discussion

In this cohort of 3,817 well-phenotyped AF patients, we identified several biomarkers associated with adverse cardiovascular events, including markers of myocardial injury (hsTropT), inflammation (IL-6), oxidative stress (GDF-15), coagulation (d-dimer), and cardiac dysfunction (NT-proBNP, IGFBP-7). The integration of these biomarkers into both traditional and machine learning-based predictive models significantly enhanced risk prediction, providing a more comprehensive assessment of adverse cardiovascular outcomes in this population.

Our analysis identified 6 biomarkers independently associated with AF-related complications and bleeding. GDF-15, a member of the TGF-β superfamily induced in cardiomyocytes, plays a significant role in oxidative stress, inflammation, cardiac injury and fibrosis ^7^. Epidemiological studies suggest that elevated GDF-15 levels increase the risk of bleeding in patients with cardiovascular diseases or AF ^8–10^. Our findings not only confirm this association but also highlight GDF-15 as a robust predictor of heart failure hospitalization, with its predicitive strength comparable to NT-proBNP and surpassing than IGFBP-7 **(Supplementary Table 5)** ^11^. IL-6, a well-established pro-inflammatory cytokine, has been linked to the pathophysiology of cardiovascular disease, particularly in AF patients^12^. Our study expands on these findings by demonstrating an association between IL-6 levels and both major and any bleeding events, suggesting that systemic inflammation may play a role in disrupting coagulation and vascular permeability. Additionally, IL-6 was significantly associated with stroke outcomes, consistent with findings from Mendelian randomization studies identifying IL-6 as a causal mediator of ischemic stroke in non-AF populations ^13^. Genetic studies also underscore the relationship between IL-6 and atherosclerosis ^14,15^, a key risk factor for stroke. These findings warrant further investigation into the causal mechanisms linking IL-6 to both ischemic stroke and bleeding, as well as its potential therapeutic implications.

Our study underscores the multifactorial nature of AF-related complications, with diverse pathophysiological pathways contributing to risk. Observational studies have shown that a multimarker approach significantly improves risk prediction in both cardiovascular disease and AF populations ^16–19^. We demonstrated that incorporating key biomarkers into prediction models significantly enhanced the discriminatory ability of Cox and most machine learning models. This supports the concept that a biomarker panel reflecting the diverse tissue changes seen in AF provides a valuable approach for comprehensive cardiovascular risk assessment. There biomarkers may offer a more detailed phenotypic profile of AF patients, and their integration into clinical decision-making could facilitate more precise management strategies, identifying those who would benefit most from intensive risk factor modification.

Targeting multiple pathophysiological systems is essential for improving outcomes in complex cardiovascular conditions. For example, the polypill strategy has shown promise in stable coronary artery disease by simultaneously addressing multiple risk factors^20^. In AF patients, targeted treatments aimed at underlying cardiovascular conditions has been shown to improve sinus rhythm maintenance in persistent AF patients^21^; however, the impact of this strategy in reducing cardiovascular outcomes remains unclear. To date, no randomized trial has specifically evaluated the effects of such a multifaceted treatment approach in AF patients. Future clinical trials should therefore investigate whether comprehensive strategies targeting inflammation, coagulation disturbances, and cardiac dysfunction can improve long-term outcomes in this high-risk population.

The CHA_2_DS_2_-VASc score is widely used tool for predicting stroke risk in AF patients^22^. However, its discriminatory performance is moderate at best^23^. Recent studies incorporating biomarkers of coagulation disturbance have shown improved stroke prediction compared to the CHA_2_DS_2_-VASc score^24,25^. In our study, the integration of NT-proBNP, hsTropT and IL-6 into the CHA_2_DS_2_-VASc score significantly improved its ability to predict all types of strokes **(Figure 3)**, despite the fact that most patients were on oral anticoagulation. While the improvement in predictive performance may appear modest, our findings suggest that biomarkers provide additional insights that could influence clinical management strategies.

We built machine learning models to leverage the entire set of clinical and biomarker variables for risk prediction. While traditional Cox regression models are limited by the number of predictors they can include without overfitting the model, machine learning models offer the advantage of capturing complex, non-linear relationships between clinical variables, biomarkers, and outcomes. In our study, we demonstrated that machine learning models, such as XGBoost and LASSO, achieved significant improvements in predictive performance when biomarkers were included. These models, coupled with biomarker panels, have the potential to help clinicians identify patients who may benefit from further investigation and treatment. Future studies should assess whether the use of machine learning-based risk models can improve the management of AF patients.

This study has several limitations. First, the models were developed using a Swiss cohort, and their performance in external validation with other AF patient cohorts remains to be determined. However, we performed repeated cross-validation on the machine learning models, supporting the robustness of the results. Second, the study population was predominantly anticoagulated, which may limit the generalizability of our results to non-anticoagulated populations.

In summary, our study highlights the complex, multifactorial nature of AF-related cardiovascular complications. We identified several biomarkers linked to diverse pathophysiological pathways – including myocardial injury, inflammation, oxidative stress, coagulation and cardiac dysfunction – that are associated with adverse cardiovascular outcomes. By integrating these biomarkers into both traditional and machine learning-based risk models, we significantly enhanced predictive accuracy, underscoring the potential clinical utility of biomarker-informed risk assessments in refining and optimizing the management in patients with AF.

## Data availability

The data supporting the findings from this study are available within the manuscript and its supplementary information. Data protection rules prohibit individual level data to be shared.

## Code availability

The code is publicly available and can be found via GitHub at https://github.com/Meyrep/biomarkers_AF_outcomes. The source code from the R-packages used in this study are freely available online (https://cran.r-project.org/).

## Author contributions

P.B.M. and D.C. conceived and planned the study. P.B.M., A.Z. and D.C. performed the technical parts and analytic approach. P.B.M. and D.C. analyzed the data and contributed to the original interpretation. P.B.M. wrote the draft manuscript and all authors discussed the results and contributed to the final manuscript.

## Methods

### Study population and procedures

We included patients with previously diagnosed AF from 2 prospective, multicenter cohort studies in Switzerland that used similar methodologies. The Basel Atrial Fibrillation (BEAT-AF) study enrolled 1,545 patients from 2010 to 2014 across 7 centers in Switzerland ^26^, and the Swiss Atrial Fibrillation (Swiss-AF) study enrolled 2,415 patients from 2014 to 2017 across 14 centers in Switzerland ^27^. Both studies had almost identical inclusion and exclusion criteria, as shown in **Supplementary Table 1**. Eligible patients had to have previously diagnosed AF. Patients who had secondary forms of AF or were unable to provide informed consent were excluded. For this analysis, we combined the BEAT-AF and Swiss-AF datasets, excluding 67 patients because of missing follow-up information and 76 patients because of missing of all biomarker values, leaving a total of 3,817 patients **(Supplementary Figure 1)**. Both studies comply with the Declaration of Helsinki, the study protocols were approved by the local ethics committees, and written informed consent was obtained from all participants.

At study enrolment and during yearly follow-up visits, trained study personnel collected information about patient demographics, risk factors, medical history, and current medical therapy using standardized case report forms. A detailed list of all variables collected are outlined in **Supplementary Table 2**. AF type was categorized according to guideline recommendations at the time of protocol development into paroxysmal, persistent, or permanent ^28^. Body mass index was calculated as weight in kilogram divided by height in meters squared. Three consecutive blood pressure measurements were obtained at study enrolment and the mean was used for all analyses. Estimated glomerular filtration rate (eGFR) was calculated using the Chronic Kidney Disease Epidemiology Collaboration (CKD-EPI) formula.

### Biomarker analyses and multiple imputation

Blood samples were drawn at baseline, immediately processed and stored at -80°C in a central biobank. We measured a biomarkers panel consisting of 12 biomarkers, selected to reflect the major disease pathways, such as vascular changes, collagen infiltration, energy metabolism, inflammatory processes, myocardial wall tension and tissue injury. These biomarkers include angiopoetin-2 (ANG-2), d-dimer, growth differentiation factor-15 (GDF-15), insulin-like growth factor-binding protein-7 (IGFBP-7), N-terminal pro-B-type natriuretic peptide (NT-proBNP), high-sensitivity troponin T (hsTropT), creatinine, cystatin C, osteopontin (OPN), high sensitive C-reactive protein (hs-CRP), interleukin-6 (IL-6), and alanine aminotransferase (ALAT). Biomarkers were analyzed centrally at Roche Diagnostics, Penzberg (Germany) on a cobas c311 or e601 by laboratory personnel blinded to clinical information under constant quality control and calibration. Most of the assays were routine products running on routine clinical analyzers. Detailed description about biomarker measurement is provided in the **Supplementary Table 3**.

To handle missing biomarker data in our dataset, we first assessed the percentage of missing values using a custom function pMiss, which calculates the proportion of missing values for each variable and each observation. The results showed that biomarkers had between 0.3% to 9.3% missing data. We then employed the mice package to visualize the missing data pattern and impute missing values using predictive mean matching (PMM) method across 5 imputations. The imputed dataset was summarized and visually inspected using density plots and strip plots to assess the distribution and consistency of the imputed values **(Supplementary Figure 2)**. This approach ensures robust handling of missing data while preserving the integrity of the biomarker dataset for subsequent statistical analyses.

### Adverse cardiovascular outcome measures

The three main outcomes of this analysis were: (1) a composite of cardiovascular death, nonfatal ischemic stroke, nonfatal systemic embolism, or nonfatal myocardial infarction, (2) heart failure hospitalization and (3) major bleeding. Secondary outcomes were the individual components of the composite outcome, as well as total stroke, myocardial infarction (MI), clinically relevant non-major (NM) bleeding, a composite of major or clinically relevant NM bleeding, and all-cause death. Definitions of all outcomes were identical in both cohorts and definitions are provided in **Supplementary Table 4**. All clinical outcomes were adjudicated.

### Development of machine learning models

To capture non-linear and complex relationships between clinical variable, biomarkers, and outcomes, different machine learning methods were applied. Specifically, 3 statistical methods – Least Absolute Shrinkage and Selection Operator (LASSO), Random Forest and Extreme Gradient Boosting for survival analysis (XGBoost)^29–31^ – were used for all outcomes. We applied three statistical methods to develop predictive models for primary and secondary outcomes. For this we included the whole sample of clinical variable (46 variables detailed in **Supplementary Table 2**) and constructed two types of models: a base model incorporating only clinical variable, and an enhanced model that included both clinical variables and all biomarkers (base model + biomarkers). The machine learning techniques used to build these models are outlined below.

#### Least Absolute Shrinkage and Selection Operator (LASSO)

LASSO was used to perform variable (feature) selection, regularization, and build parsimonious models for the primary and secondary outcomes. In brief, LASSO is a regularized regression analysis that simultaneously performs variable selection and regularization using machine learning algorithms. First, the feature matrix was prepared by converting the dataset into a model matrix excluding the intercept column. The response variable was set as the outcome being studied. LASSO logistic regression was then performed using the *cv.glmnet* function from the *glmnet* package, with 10-fold cross-validation to optimize the model’s regularization parameter (lambda). The optimal lambda was determined by the value that minimized the cross-validated error (*lambda.min*). The resulting model coefficients were extracted, and non-zero coefficients were identified as the selected variables.

#### Random Forest

Random Forest, an ensemble learning method that enhances predictive accuracy by combining multiple decision trees, was used to develop models for predicting primary and secondary outcomes. Each variable was considered as a potential predictor, and the algorithm built an ensemble of decision trees, each trained on different subsets of variables. The final prediction was derived from the average of individual tree predictions. The Random Forest models were constructed using the *randomForest* package in R, with reproducibility ensured by setting a random seed. The models were trained with default settings. Predictive probabilities for the composite outcome were generated using the trained model, and model accuracy was evaluated using the *confusionMatrix* function. Additionally, model performance was assessed through 10-fold cross-validation across 100 decision trees.

#### Extreme Gradient Boosting (XGBoost)

The XGBoost algorithm builds a series of models for the outcome being studied, each focusing on correcting residuals (“boosting”) of the combined predictions of the models built so far. Specifically, each new model aims to capture the relationships in the data that were not well represented by the previous models. To develop predictive models using the XGBoost algorithm, we transformed outcome variables to a binary numeric format. The XGBoost model was trained on the dataset with a random seed set for reproducibility, using the *xgboost* package in R. The model was configured with a binary logistic objective and trained over 10 boosting rounds. Post-training, variable importance was assessed using the *xgb.importance* function, which ranked the features based on their contribution to the model. True outcomes were used to evaluate the model’s performance, allowing for a robust assessment of its predictive capabilities.

## Statistical analyses

The normality of distribution of each biomarker was assessed through visual inspection of histograms. Spearman rank correlations were applied to evaluate interrelationships between biomarkers. All biomarkers were log-transformed to improve the normality of the distribution. Cox proportional hazard models were constructed to assess hazard ratios (HR) and 95% confidence intervals (CI) for the main and secondary outcomes. To provide a unit-independent comparison between log-transformed biomarkers, HRs were standardized, representing the effect per 1 standard deviation (SD) increase. The initial model was adjusted for age and sex and the multivariable model was adjusted for a prespecified list of covariates, including age, sex, body mass index, current smoker, history of hypertension, history of diabetes, prior stroke, history of heart failure, chronic kidney disease, and coronary artery disease. First separate models were constructed for each biomarker. We then constructed a combined multivariable model including all biomarkers in a single model and performed backward selection of the biomarkers using the Akaike information criterion (AIC) to exclude biomarkers. Standardized HRs were plotted using coefficient plots and against the -log10 p-value using volcano plots. In addition, we measured each variable’s relative importance in the models by calculating the partial χ2 statistic minus the predictor degrees of freedom. We then calculated the Area Under the Curve (AUC) of the Receiver Operating Characteristic (ROC) curve with corresponding 95% CI for each Cox model with and without key biomarkers and compared them using DeLong’s test. Finally, we used the CHA_2_DS_2_-VASc score as a base Cox model for ischemic stroke and all strokes and used backward selection to optimize the model with biomarkers.

We constructed predictive machine learning models using the entire sample of clinical variables **(Supplementary Table 2)** to construct 2 types of models: a base model that only includes clinical variables, and an enhanced model incorporating both clinical variables and all biomarkers. The model’s discriminative ability was evaluated using AUC and corresponding 95% CI. AUC between base model and base model + biomarkers were compared using DeLong’s tests. All statistical analyses were performed using R version 4.3.1. A P-value <0.05 was considered statistically significant.

