## Supplement for "Biomarker panels for improved risk prediction and enhanced biological insights in patients with atrial fibrillation"

**Supplementary Table 1.** Description of cohorts

**Supplementary Table 2.** Variables collected at baseline and included in the machine learning models

**Biomarker measurements Table 3.** Description of biomarker measurements

**Supplementary Table 4.** Definition of major adverse cardiac events

**Supplementary Table 5.** Biomarkers and risk of composite of cardiovascular death, nonfatal ischemic stroke, nonfatal systemic embolism and nonfatal myocardial infarction

**Supplementary Table 6.** Biomarkers and risk of heart failure hospitalization

**Supplementary Table 7.** Biomarkers and risk of major bleeding

**Supplementary Table 8.** Biomarkers and risk of ischemic stroke

**Supplementary Table 9.** Biomarkers and risk of all strokes

**Supplementary Table 10.** Biomarkers and risk of myocardial infarction

**Supplementary Table 11.** Biomarkers and risk of cardiovascular death

**Supplementary Table 12.** Biomarkers and risk of all-cause death

**Supplementary Table 13.** Biomarkers and risk of any bleeding

**Supplementary Table 14.** Biomarkers and risk of clinically relevant non-major bleeding

**Supplementary Figure 1.** Flow diagram of the study

**Supplementary Figure 2.** Missing pattern and multiple imputation of biomarkers

**Supplementary Figure 3.** Spearman rank correlations of biomarkers

**Supplementary Figure 4.** Risk of adverse cardiovascular outcomes by biomarkers

**Supplementary Figure 5.** Predictive performance of Cox and machine learning models for outcomes with and without biomarkers

**Supplementary Table 1. Description of cohorts**

| **BEAT-AF** | **Swiss-AF** |
| --- | --- |
| N=1,546 | N=2,415 |
| **Inclusion criteria** | |
| Documented (by electrocardiogram [ECG], rhythm strip or device interrogation) paroxysmal AF, persistent AF or permanent AF | Documented (by ECG, rhythm strip or device interrogation) paroxysmal AF, persistent AF or permanent AF  Age ≥65 years |
| **Exclusion criteria** | |
| Any acute illness within the last 4 weeks. These patients could be enrolled after stabilization of their acute episode | Any acute illness within the last 4 weeks. These patients could be enrolled after stabilization of their acute episode |
| Patients who exclusively have short episodes of reversible forms of AF (e.g. AF post cardiac surgery, thyrotoxicosis) | Patients who exclusively have short episodes of reversible forms of AF (e.g. AF post cardiac surgery, thyrotoxicosis) |
| Inability to sign informed consent | Inability to sign informed consent |
|  | Participating in BEAT-AF |
| **Participating Study Centers** | |
| University Hospital Basel  Cantonal Hospital St. Gallen  Cantonal Hospital Bellinzona  Cantonal Hospital Lugano  Hospital Rheinfelden  University Hospital Zürich  University Hospital Lausanne  Cantonal Hospital Lucerne  University Hospital Geneva | University Hospital Basel  Cantonal Hospital St. Gallen  Cantonal Hospital Bellinzona  Cantonal Hospital Lugano  University Hospital Lausanne  Cantonal Hospital Lucerne  University Hospital Geneva  Cantonal Hospital Baden  University Hospital Berne  Cardiocentro Ticino, Lugano  Cantonal Hospital Fribourg  Hospital St. Anna, Lucerne  Stadtspital Triemli, Zürich  Cantonal Hospital Solothurn |

**Supplementary Table 2. Variables collected at baseline and included in the machine learning models**

| **Variable** | **Description** |
| --- | --- |
| Age | years |
| Sex | Male, female |
| BMI | kg/m^2^ |
| Smoking | Yes=active or No=Past or never |
| Systolic blood pressure | mmHg |
| Diabetes | Yes or No |
| Prior stroke or TIA | Yes or No |
| Heart failure | Yes or No |
| Renal failure | Yes or No |
| Coronary artery disease | Yes or No |
| Study center | University Hospital Basel and Basel University, University Hospital Bern, Stadtspital Triemli Zurich, Kantonspital Baden, Cardiocentro Lugano, Kantonsspital St. Gallen, Hôpital Cantonal Fribourg, Luzerner Kantonsspital, Ente Ospedaliero Cantonale Lugano, University Hospital Geneva, University Hospital Lausanne, Bürgerspital Solothurn, Ente Ospedaliero Cantonale Bellinzona, University of Zurich/University Hospital Zurich, Hospital Rheinfelden, Hospital St. Anna, Lucerne |
| Ethnicity | Central Europe, Southern Europe, Northern Europe, Eastern Europe, Central/South America, North America, Other |
| AF type | Paroxysmal, persistent, permanent |
| Beer drinker | >6/day, 4-5/day, 2-3/day, 1/day, 5-6/week, 2-4/week, 1/week, 1-3/month, never or <1/month |
| Red wine drinker | >6/day, 4-5/day, 2-3/day, 1/day, 5-6/week, 2-4/week, 1/week, 1-3/month, never or <1/month |
| White wine drinker | >6/day, 4-5/day, 2-3/day, 1/day, 5-6/week, 2-4/week, 1/week, 1-3/month, never or <1/month |
| Liquor drinker | >6/day, 4-5/day, 2-3/day, 1/day, 5-6/week, 2-4/week, 1/week, 1-3/month, never or <1/month |
| Atrial fibrillation duration | Permanent, days, hours, minutes, no more |
| Nr. of atrial fibrillation episodes | Permanent, >1/week, 1/week, <1/week but >1/month, <1/month, no more |
| Atrial flutter | Yes or No |
| Aspirin | Yes or No |
| Antiplatelet therapy | Yes or No |
| Ticagrelor | Yes or No |
| Ilicit drugs | Yes or No |
| Cancer | Yes or No |
| VTE | Yes or No |
| Paternal AF history | Father with a history of AF, Yes or No |
| Brother AF history | Brother with a history of AF , Yes or No |
| Maternal AF history | Mother with a history of AF, Yes or No |
| Sister AF history | Sister with a history of AF , Yes or No |
| Family history of hypertension | Yes or No |
| Family history of diabetes | Yes or No |
| Family history of obesity | Yes or No |
| Family history of coronary artery disease | Yes or No |
| Rhythm on ECG at inclusion | Atrial fibrillation, atrial flutter, sinus rhythm, other |
| Hyperthyroidism | Yes or No |
| Hypothyroidism | Yes or No |
| Prior CABG | Yes or No |
| Height | cm |
| Weight | kg |
| Heart rate | bpm |
| Diastolic blood pressure | mmHg |
| Hypertension | Yes or No |
| Peripheral artery disease | Yes or No |
| Prior myocardial infarction | Yes or No |
| Systemic embolism | Yes or No |
| Prior major bleeding | Yes or No |

**Supplementary Table 3. Description of biomarker measurements**

| **Biomarker** | **Methods** |
| --- | --- |
| D-dimer | D-dimer values were analyzed via a commercial Tina-quant D-Dimer Gen.2 test (Roche Diagnostics, Mannheim, Germany) on a cobas C311 analyzer (Roche) according to the manufacturer’s instructions. Values are provided in ug/mL. The limit of detection (LoD) was 0.150 ug/ml. |
| IL-6 | The commercial Elecsys IL-6 assay (cobas e601, Roche Diagnostics, Mannheim, Germany) was used to measure plasma levels of IL-6. The minimal determined IL-6 was 1.5 pg/ml (LoD) |
| NT-proBNP | NT-proBNP was determined using the commercial Roche Elecsys proBNP II IVD on a cobas e601 (measuring range 10–35000ng/L) with a coefficient of variation of 2.45% for the lower control measured (mean 133.1ng/L). The assays applied are based on the Elecsys electro-chemiluminescence technology. |
| IGFBP-7 | IGFBP-7 levels were measured with a precommercial Elecsys assay using an automated cobas electrochemiluminescence immunoassay analyzer e601 (Roche Diagnostics, Mannheim, Germany) For IGFBP-7, the limit of detection was 0.01 ng/mL, and the within-run precision coefficient of variation was 2%. |
| Cystatin C | Cystatin C levels were determined with the commercial Tina-quant Cystatin C Gen.2 assay (cobas c 311; Roche Diagnostics, Mannheim, Germany) with a lower limit of detection of 0.4 mg/L. |
| Hs-CRP | The commercial Cardiac C-Reactive Protein (Latex) High Sensitive assay was used (cobas c 311, Roche Diagnostics, Mannheim, Germany) to measure plasma levels of hs-CRP. The lower limit of detection was 0.15 mg/L. |
| OPN | OPN levels were measured with a precommercial Elecsys assay using an automated cobas electrochemiluminescence immunoassay analyzer e601 (Roche Diagnostics, Mannheim, Germany). For OPN, the limit of detection was 0.01 ng/mL. |
| GDF-15 | Growth differentiation factor-15 (GDF-15), was determined by commercially available Elecsys assay on a e601 (Elecsys®; Roche Diagnostics, Mannheim, Germany). LoD was 22pg/mL. |
| hsTropT | Levels of hs-Troponin T (TNTHS) were determined by commercially available Elecsys assay on a e601 (oche Diagnostics, Mannheim, Germany). LoD was 3 ng/L. |
| ANG-2 | Ang2 levels were measured with a precommercial Elecsys assay using an automated cobas electrochemiluminescence immunoassay analyzer e601 (Roche Diagnostics, Mannheim, Germany). LoD was 0.03 ng/mL. |
| ALAT | ALAT levels were determined with the commercially available ALTL assay (cobas c 311; Roche Diagnostics, Mannheim, Germany) with a LoD of 5 U/L (0.08 μkat/L). |

**Supplementary Table 4. Definition of major adverse cardiac events**

Major bleeding

Major bleeding is defined according to the International Society on Thrombosis and Haemostasis criteria, as clinically overt bleeding with a fatal outcome, a reduction in haemoglobin level of ≥20 g/l within 7 days, transfusion of at least two units of blood, or symptomatic bleeding in a critical area or organ (intracranial, intraspinal, intraocular, pericardial, intra-articular, intramuscular with compartment syndrome, retroperitoneal).(1)

Clinically relevant non-major bleeding

CRNMB is defined as bleeding not fulfilling the ISTH criteria, but that was clinically overt and led to hospitalization, change of antithrombotic therapy, or necessitated a medical or surgical intervention.(2)

Stroke

Stroke is defined as an acute focal neurological deficit of vascular origin, with evidence of focal infarction confirmed by imaging (computed tomography or cMRI) or autopsy. Stroke is categorised as ischaemic, haemorrhagic or of unknown cause (based on computed tomography, cMRI, autopsy and other ancillary investigations). Ischaemic strokes are further classified according to the TOAST classification.(3) Fatal stroke is defined as death from any cause within 30 days after stroke.

Myocardial infarction

Myocardial infarction is defined as rise and/or fall of cardiac troponin with at least one value above the 99th percentile of the upper reference limit in a clinical setting consistent with myocardial ischaemia, and with at least one of the following: symptoms of ischaemia, new significant ST-T changes or new left bundle-branch block on ECG, development of pathological Q waves in the ECG, imaging evidence of new loss of viable myocardium or new regional wall motion abnormality, identification of an intracoronary thrombus by angiography or autopsy.(4)

Death

The cause of death is classified as being of cardiac, stroke, any cardiovascular, bleeding, non-cardiovascular or unknown aetiology.

**Supplementary Table 4. Biomarkers and risk of composite of cardiovascular death, nonfatal ischemic stroke, nonfatal systemic embolism and nonfatal myocardial infarction**

|  | **Age and sex adjusted model** | | **Multivariable model** | | **Combined model**  **(backward selection)** | |
| --- | --- | --- | --- | --- | --- | --- |
| **Biomarker** | **HR (95% CI)*** | **P value** | **HR (95% CI)** | **P value** | **HR (95% CI)** | **P value** |
| ANG-2 | 1.48 (1.39-1.58) | <2x10^-16^ | 1.35 (1.26-1.45) | <2x10^-16^ | 1.07 (0.98-1.17) | 0.141978 |
| eGFR | 0.71 (0.65-0.77) | 2.9x10^-15^ | 0.79 (0.70-0.88) | 1.5x10^-5^ | - | - |
| Cystatin C | 1.43 (1.36-1.51) | <2x10^-16^ | 1.31 (1.23-1.40) | <2x10^-16^ | - | - |
| D-dimer | 1.23 (1.17-1.30) | 4.4x10^-16^ | 1.18 (1.12-1.24) | 1.8x10^-9^ | 1.09 (1.02-1.14) | 0.010065 |
| ALAT | 0.87 (0.81-0.95) | 0.00104 | 0.88 (0.81-0.95) | 0.00146 | 0.87 (0.81-0.94) | 0.000476 |
| GDF-15 | 1.79 (1.67-1.92) | <2x10^-16^ | 1.58 (1.45-1.72) | <2x10^-16^ | 1.22 (1.11-1.35) | 7.5x10^-5^ |
| Hs-CRP | 1.21 (1.14-1.29) | 1.9x10^-9^ | 1.15 (1.08-1.23) | 3.4x10^-5^ | - | - |
| IGFBP-7 | 1.54 (1.44-1.64) | <2x10^-16^ | 1.39 (1.29-1.50) | <2x10^-16^ | - | - |
| IL-6 | 1.41 (1.34-1.49) | <2x10^-16^ | 1.31 (1.24-1.40) | <2x10^-16^ | 1.12 (1.04-1.21) | 0.002527 |
| NT-proBNP | 1.91 (1.75-2.09) | <2x10^-16^ | 1.69 (1.53-1.86) | <2x10^-16^ | 1.30 (1.15-1.47) | 2.1x10^-5^ |
| OPN | 1.49 (1.41-1.58) | <2x10^-16^ | 1.39 (1.29-1.49) | <2x10^-16^ | - | - |
| hsTropT | 1.57 (1.48-1.66) | <2x10^-16^ | 1.47 (1.38-1.57) | <2x10^-16^ | 1.21 (1.11-1.32) | 8.1x10^-6^ |
| *Hazard ratios are standardized per 1-SD increase in biomarker. Multivariable models are adjusted for additional factors controlled for body mass index, current smoker, systolic blood pressure, history of diabetes, prior stroke or TIA, history of heart failure, chronic kidney disease and coronary artery disease. The combined model is adjusted for covariates from the multivariable model plus all significant biomarkers after backward selection based on the variables p-values. | | | | | | |

**Supplementary Table 5. Biomarkers and risk of heart failure hospitalization**

|  | **Age and sex adjusted model** | | **Multivariable model** | | **Combined model**  **(backward selection)** | |
| --- | --- | --- | --- | --- | --- | --- |
| **Biomarker** | **HR (95% CI)*** | **P value** | **HR (95% CI)** | **P value** | **HR (95% CI)** | **P value** |
| ANG-2 | 1.61 (1.51-1.72) | <2x10^-16^ | 1.41 (1.31-1.51) | <2x10^-16^ | - | - |
| eGFR | 0.68 (0.62-0.74) | <2x10^-16^ | 0.76 (0.68-0.84) | 7.3x10^-7^ | - | - |
| Cystatin C | 1.53 (1.46-1.61) | <2x10^-16^ | 1.34 (1.26-1.43) | <2x10^-16^ | - | - |
| D-dimer | 1.22 (1.15-1.28) | 3.3x10^-13^ | 1.13 (1.07-1.20) | 1.1x10^-5^ | - | - |
| ALAT | 0.92 (0.85-1.00) | 0.06 | 0.93 (0.85-1.01) | 0.07 | 0.91 (0.84-0.99) | 0.03060 |
| GDF-15 | 2.14 (1.99-2.30) | <2x10^-16^ | 1.86 (1.70-2.03) | <2x10^-16^ | 1.49 (1.33-1.66) | 6.2x10^-12^ |
| Hs-CRP | 1.28 (1.20-1.36) | 1.1x10^-13^ | 1.16 (1.08-1.24) | 2.6x10^-5^ | - | - |
| IGFBP-7 | 1.76 (1.65-1.88) | <2x10^-16^ | 1.56 (1.44-1.68) | <2x10^-16^ | 1.12 (1.01-1.24) | 0.03576 |
| IL-6 | 1.44 (1.35-1.52) | <2x10^-16^ | 1.28 (1.20-1.37) | 1.4x10^-13^ | - | - |
| NT-proBNP | 2.36 (2.14-2.60) | <2x10^-16^ | 1.99 (1.79-2.21) | <2x10^-16^ | 1.55 (1.37-1.74) | 1.9x10^-12^ |
| OPN | 1.64 (1.55-1.74) | <2x10^-16^ | 1.40 (1.30-1.51) | <2x10^-16^ | - | - |
| hsTropT | 1.67 (1.58-1.76) | <2x10^-16^ | 1.52 (1.42-1.62) | <2x10^-16^ | 1.22 (1.11-1.34) | 2.1x10^-5^ |
| *Hazard ratios are standardized per 1-SD increase in biomarker. Multivariable models are adjusted for additional factors controlled for body mass index, current smoker, systolic blood pressure, history of diabetes, prior stroke or TIA, history of heart failure, chronic kidney disease and coronary artery disease. The combined model is adjusted for covariates from the multivariable model plus all significant biomarkers after backward selection based on Akaike Information Criterion (AIC). | | | | | | |

**Supplementary Table 6. Biomarkers and risk of major bleeding**

|  | **Age and sex adjusted model** | | **Multivariable model** | | **Combined model**  **(backward selection)** | |
| --- | --- | --- | --- | --- | --- | --- |
| **Biomarker** | **HR (95% CI)*** | **P value** | **HR (95% CI)** | **P value** | **HR (95% CI)** | **P value** |
| ANG-2 | 1.22 (1.11-1.34) | 3.8x10^-5^ | 1.16 (1.05-1.28) | 0.00399 | - | - |
| eGFR | 0.80 (0.71-0.90) | 0.000339 | 0.95 (0.82-1.11) | 0.52 | - | - |
| Cystatin C | 1.32 (1.21-1.43) | 5.6x10^-11^ | 1.24 (1.12-1.37) | 5.6x10^-5^ | - | - |
| D-dimer | 1.12 (1.04-1.21) | 0.0043 | 1.08 (0.99-1.18) | 0.06 | - | - |
| ALAT | 0.93 (0.84-1.04) | 0.20 | 0.95 (0.86-1.06) | 0.34 | - | - |
| GDF-15 | 1.53 (1.38-1.69) | 1.3x10^-15^ | 1.46 (1.29-1.65) | 1.4x10^-9^ | 1.30 (1.12-1.52) | 0.000644 |
| Hs-CRP | 1.12 (1.03-1.23) | 0.012 | 1.14 (0.99-1.20) | 0.061 | - | - |
| IGFBP-7 | 1.42 (1.29-1.56) | 5.3x10^-13^ | 1.32 (1.19-1.47) | 3.6x10^-7^ | 1.14 (1.01-1.30) | 0.048090 |
| IL-6 | 1.28 (1.18-1.40) | 7.4x10^-9^ | 1.23 (1.13-1.35) | 3.2x10^-6^ | 1.11 (1.01-1.23) | 0.039263 |
| NT-proBNP | 1.37 (1.21-1.54) | 2.7x10^-7^ | 1.26 (1.11-1.43) | 0.00043 | - | - |
| OPN | 1.37 (1.25-1.50) | 3.3x10^-12^ | 1.28 (1.16-1.43) | 3.6x10^-6^ | - | - |
| hsTropT | 1.39 (1.27-1.52) | 1.2x10^-12^ | 1.31 (1.19-1.45) | 1.2x10^-7^ | 1.16 (1.03-1.31) | 0.014368 |
| *Hazard ratios are standardized per 1-SD increase in biomarker. Multivariable models are adjusted for additional factors controlled for body mass index, current smoker, systolic blood pressure, history of diabetes, prior stroke or TIA, history of heart failure, chronic kidney disease and coronary artery disease. The combined model is adjusted for covariates from the multivariable model plus all significant biomarkers after backward selection based on Akaike Information Criterion (AIC). | | | | | | |

**Supplementary Table 7. Biomarkers and risk of ischemic stroke**

|  | **Age and sex adjusted model** | | **Multivariable model** | | **Combined model**  **(backward selection)** | |
| --- | --- | --- | --- | --- | --- | --- |
| **Biomarker** | **HR (95% CI)*** | **P value** | **HR (95% CI)** | **P value** | **HR (95% CI)** | **P value** |
| ANG-2 | 1.27 (1.11-1.47) | 0.00072 | 1.21 (1.04-1.41) | 0.0116 | - | - |
| eGFR | 0.97 (0.80-1.18) | 0.77 | 1.09 (0.86-1.33) | 0.43 | - | - |
| Cystatin C | 1.08 (0.92-1.27) | 0.34 | 1.02 (0.84-1.23) | 0.88 | - | - |
| D-dimer | 1.07 (0.94-1.22) | 0.29 | 1.08 (0.94-1.23) | 0.28 | - | - |
| ALAT | 0.91 (0.78-1.06) | 0.24 | 0.91 (0.78-1.06) | 0.23 | 0.87 (0.74-1.02) | 0.0891 |
| GDF-15 | 1.18 (0.99-1.39) | 0.06 | 1.06 (0.87-1.30) | 0.56 | - | - |
| Hs-CRP | 1.09 (0.95-1.25) | 0.24 | 1.08 (0.94-1.24) | 0.29 | - | - |
| IGFBP-7 | 1.13 (0.97-1.33) | 0.13 | 1.04 (0.87-1.25) | 0.65 | - | - |
| IL-6 | 1.19 (1.04-1.36) | 0.013 | 1.15 (0.99-1.32) | 0.07 | - | - |
| NT-proBNP | 1.52 (1.27-1.82) | 3.6x10^-6^ | 1.49 (1.23-1.81) | 5.6x10^-5^ | 1.61 (1.30-1.98) | 8.4x10^-6^ |
| OPN | 1.02 (0.86-1.21) | 0.81 | 0.95 (0.77-1.15) | 0.58 | 0.82 (0.66-1.01) | 0.0565 |
| hsTropT | 1.20 (1.03-1.40) | 0.023 | 1.14 (0.95-1.36) | 0.15 | - | - |
| *Hazard ratios are standardized per 1-SD increase in biomarker. Multivariable models are adjusted for additional factors controlled for body mass index, current smoker, systolic blood pressure, history of diabetes, prior stroke or TIA, history of heart failure, chronic kidney disease and coronary artery disease. The combined model is adjusted for covariates from the multivariable model plus all significant biomarkers after backward selection based on Akaike Information Criterion (AIC). | | | | | | |

**Supplementary Table 8. Biomarkers and risk of all strokes**

|  | **Age and sex adjusted model** | | **Multivariable model** | | **Combined model**  **(backward selection)** | |
| --- | --- | --- | --- | --- | --- | --- |
| **Biomarker** | **HR (95% CI)*** | **P value** | **HR (95% CI)** | **P value** | **HR (95% CI)** | **P value** |
| ANG-2 | 1.25 (1.10-1.42) | 0.00055 | 1.20 (1.05-1.37) | 0.0072 | - | - |
| eGFR | 0.98 (0.83-1.16) | 0.83 | 1.07 (0.89-1.29) | 0.49 | - | - |
| Cystatin C | 1.09 (0.95-1.25) | 0.22 | 1.06 (0.89-1.25) | 0.52 | - | - |
| D-dimer | 1.10 (0.98-1.22) | 0.11 | 1.10 (0.98-1.24) | 0.09 | - | - |
| ALAT | 0.93 (0.81-1.07) | 0.31 | 0.92 (0.80-1.06) | 0.26 | 0.90 (0.78-1.03) | 0.12 |
| GDF-15 | 1.16 (0.99-1.35) | 0.051 | 1.09 (0.91-1.31) | 0.33 | - | - |
| Hs-CRP | 1.09 (0.97-1.24) | 0.15 | 1.09 (0.97-1.24) | 0.16 | - | - |
| IGFBP-7 | 1.14 (0.99-1.32) | 0.07 | 1.08 (0.92-1.27) | 0.35 | - | - |
| IL-6 | 1.23 (1.09-1.38) | 0.00054 | 1.20 (1.06-1.36) | 0.004 | 1.16 (1.01-1.32) | 0.0334 |
| NT-proBNP | 1.54 (1.31-1.80) | 1.0x10^-7^ | 1.53 (1.28-1.82) | 1.8x10^-6^ | 1.55 (1.29-1.87) | 4.1x10^-6^ |
| OPN | 1.08 (0.93-1.25) | 0.31 | 1.04 (0.88-1.24) | 0.63 | 0.86 (0.72-1.04) | 0.12 |
| hsTropT | 1.23 (1.08-1.41) | 0.0025 | 1.20 (1.03-1.39) | 0.017 | - | - |
| *Hazard ratios are standardized per 1-SD increase in biomarker. Multivariable models are adjusted for additional factors controlled for body mass index, current smoker, systolic blood pressure, history of diabetes, prior stroke or TIA, history of heart failure, chronic kidney disease and coronary artery disease. The combined model is adjusted for covariates from the multivariable model plus all significant biomarkers after backward selection based on Akaike Information Criterion (AIC). | | | | | | |

**Supplementary Table 9. Biomarkers and risk of myocardial infarction**

|  | **Age and sex adjusted model** | | **Multivariable model** | | **Combined model**  **(backward selection)** | |
| --- | --- | --- | --- | --- | --- | --- |
| **Biomarker** | **HR (95% CI)*** | **P value** | **HR (95% CI)** | **P value** | **HR (95% CI)** | **P value** |
| ANG-2 | 1.27 (1.09-1.48) | 0.0025 | 1.14 (0.97-1.34) | 0.11 | - | - |
| eGFR | 0.82 (0.67-1.01) | 0.06 | 0.84 (0.66-1.08) | 0.17 | - | - |
| Cystatin C | 1.32 (1.15-1.51) | 7.5x10^-5^ | 1.14 (0.95-1.37) | 0.15 | - | - |
| D-dimer | 1.15 (1.01-1.31) | 0.036 | 1.09 (0.95-1.25) | 0.22 | - | - |
| ALAT | 0.97 (0.82-1.15) | 0.75 | 0.93 (0.79-1.11) | 0.44 | - | - |
| GDF-15 | 1.56 (1.33-1.84) | 9.7x10^-8^ | 1.29 (1.05-1.58) | 0.0156 | - | - |
| Hs-CRP | 1.17 (1.02-1.36) | 0.031 | 1.09 (0.94-1.27) | 0.27 | - | - |
| IGFBP-7 | 1.30 (1.10-1.53) | 0.00179 | 1.13 (0.94-1.36) | 0.21 | - | - |
| IL-6 | 1.35 (1.19-1.54) | 7.3x10^-6^ | 1.24 (1.07-1.44) | 0.0037 | 1.19 (1.02-1.39) | 0.03068 |
| NT-proBNP | 1.46 (1.20-1.77) | 0.00012 | 1.27 (1.03-1.57) | 0.0257 | - | - |
| OPN | 1.35 (1.17-1.56) | 5.3x10^-5^ | 1.18 (0.98-1.42) | 0.07 | - | - |
| hsTropT | 1.43 (1.24-1.64) | 4.8x10^-7^ | 1.29 (1.08-1.54) | 0.0052 | 1.21 (1.01-1.45) | 0.04806 |
| *Hazard ratios are standardized per 1-SD increase in biomarker. Multivariable models are adjusted for additional factors controlled for body mass index, current smoker, systolic blood pressure, history of diabetes, prior stroke or TIA, history of heart failure, chronic kidney disease and coronary artery disease. The combined model is adjusted for covariates from the multivariable model plus all significant biomarkers after backward selection based on Akaike Information Criterion (AIC). | | | | | | |

**Supplementary Table 10. Biomarkers and risk of cardiovascular death**

|  | **Age and sex adjusted model** | | **Multivariable model** | | **Combined model**  **(backward selection)** | |
| --- | --- | --- | --- | --- | --- | --- |
| **Biomarker** | **HR (95% CI)*** | **P value** | **HR (95% CI)** | **P value** | **HR (95% CI)** | **P value** |
| ANG-2 | 1.62 (1.50-1.74) | <2x10^-16^ | 1.44 (1.33-1.56) | <2x10^-16^ | - | - |
| eGFR | 0.64 (0.58-0.70) | <2x10^-16^ | 0.71 (0.62-0.80) | 9.8x10^-8^ | - | - |
| Cystatin C | 1.55 (1.47-1.63) | <2x10^-16^ | 1.41 (1.33-1.51) | <2x10^-16^ | - | - |
| D-dimer | 1.27 (1.20-1.34) | <2x10^-16^ | 1.19 (1.12-1.27) | 6.8x10^-9^ | 1.06 (0.99-1.14) | 0.078298 |
| ALAT | 0.82 (0.74-0.90) | 5.5x10^-5^ | 0.83 (0.75-0.92) | 0.00022 | 0.83 (0.76-0.92) | 0.000150 |
| GDF-15 | 2.14 (1.97-2.32) | <2x10^-16^ | 1.86 (1.69-2.06) | <2x10^-16^ | 1.35 (1.21-1.52) | 1.6x10^-7^ |
| Hs-CRP | 1.27 (1.18-1.37) | 1.4x10^-10^ | 1.19 (1.10-1.28) | 1.1x10^-5^ | - | - |
| IGFBP-7 | 1.73 (1.61-1.86) | <2x10^-16^ | 1.55 (1.42-1.68) | <2x10^-16^ | - | - |
| IL-6 | 1.51 (1.42-1.60) | <2x10^-16^ | 1.39 (1.29-1.48) | <2x10^-16^ | 1.16 (1.07-1.26) | 0.000643 |
| NT-proBNP | 2.41 (2.16-2.70) | <2x10^-16^ | 2.08 (1.84-2.36) | <2x10^-16^ | 1.48 (1.30-1.70) | 9.7x10^-9^ |
| OPN | 1.67 (1.57-1.78) | <2x10^-16^ | 1.57 (1.44-1.70) | <2x10^-16^ | - | - |
| hsTropT | 1.76 (1.66-1.87) | <2x10^-16^ | 1.65 (1.54-1.78) | <2x10^-16^ | 1.30 (1.18-1.44) | 7.5x10^-8^ |
| *Hazard ratios are standardized per 1-SD increase in biomarker. Multivariable models are adjusted for additional factors controlled for body mass index, current smoker, systolic blood pressure, history of diabetes, prior stroke or TIA, history of heart failure, chronic kidney disease and coronary artery disease. The combined model is adjusted for covariates from the multivariable model plus all significant biomarkers after backward selection based on Akaike Information Criterion (AIC). | | | | | | |

**Supplementary Table 11. Biomarkers and risk of all-cause death**

|  | **Age and sex adjusted model** | | **Multivariable model** | | **Combined model**  **(backward selection)** | |
| --- | --- | --- | --- | --- | --- | --- |
| **Biomarker** | **HR (95% CI)*** | **P value** | **HR (95% CI)** | **P value** | **HR (95% CI)** | **P value** |
| ANG-2 | 1.55 (1.46-1.65) | <2x10^-16^ | 1.40 (1.31-1.49) | <2x10^-16^ | - | - |
| eGFR | 0.63 (0.58-0.68) | <2x10^-16^ | 0.67 (0.61-0.74) | 8.7x10^-16^ | - | - |
| Cystatin C | 1.56 (1.50-1.62) | <2x10^-16^ | 1.45 (1.38-1.53) | <2x10^-16^ | - | - |
| D-dimer | 1.27 (1.22-1.33) | <2x10^-16^ | 1.20 (1.14-1.26) | 5.1x10^-14^ | 1.07 (1.01-1.13) | 0.018502 |
| ALAT | 0.86 (0.80-0.93) | 0.00012 | 0.88 (0.81-0.95) | 0.00088 | 0.88 (0.82-0.95) | 0.001143 |
| GDF-15 | 2.17 (2.04-2.32) | <2x10^-16^ | 1.95 (1.80-2.10) | <2x10^-16^ | 1.42 (1.28-1.57) | 1.3x10^-11^ |
| Hs-CRP | 1.33 (1.25-1.40) | <2x10^-16^ | 1.24 (1.17-1.32) | 8.1x10^-13^ | - | - |
| IGFBP-7 | 1.76 (1.66-1.86) | <2x10^-16^ | 1.62 (1.52-1.73) | <2x10^-16^ | 1.12 (1.02-1.23) | 0.014693 |
| IL-6 | 1.52 (1.45-1.60) | <2x10^-16^ | 1.41 (1.34-1.49) | <2x10^-16^ | 1.19 (1.12-1.28) | 1.5x10^-7^ |
| NT-proBNP | 2.15 (1.96-2.34) | <2x10^-16^ | 1.88 (1.70-2.07) | <2x10^-16^ | 1.25 (1.12-1.39) | 7.8x10^-5^ |
| OPN | 1.68 (1.60-1.76) | <2x10^-16^ | 1.59 (1.50-1.69) | <2x10^-16^ | 1.02 (0.93-1.11) | 0.72 |
| hsTropT | 1.71 (1.62-1.79) | <2x10^-16^ | 1.60 (1.50-1.69) | <2x10^-16^ | 1.22 (1.12-1.32) | 3.2x10^-6^ |
| *Hazard ratios are standardized per 1-SD increase in biomarker. Multivariable models are adjusted for additional factors controlled for body mass index, current smoker, systolic blood pressure, history of diabetes, prior stroke or TIA, history of heart failure, chronic kidney disease and coronary artery disease. The combined model is adjusted for covariates from the multivariable model plus all significant biomarkers after backward selection based on Akaike Information Criterion (AIC). | | | | | | |

**Supplementary Table 12. Biomarkers and risk of any bleeding**

|  | **Age and sex adjusted model** | | **Multivariable model** | | **Combined model**  **(backward selection)** | |
| --- | --- | --- | --- | --- | --- | --- |
| **Biomarker** | **HR (95% CI)*** | **P value** | **HR (95% CI)** | **P value** | **HR (95% CI)** | **P value** |
| ANG-2 | 1.20 (1.13-1.28) | 4.0x10^-8^ | 1.15 (1.07-1.23) | 9.3x10^-5^ | - | - |
| eGFR | 0.90 (0.82-0.98) | 0.0170 | 0.94 (0.84-1.04) | 0.23 | - | - |
| Cystatin C | 1.21 (1.14-1.29) | 4.1x10^-9^ | 1.16 (1.07-1.25) | 0.00028 | - | - |
| D-dimer | 1.10 (1.04-1.17) | 0.0011 | 1.08 (1.02-1.15) | 0.01025 | - | - |
| ALAT | 0.97 (0.91-1.04) | 0.46 | 0.97 (0.91-1.04) | 0.41 | - | - |
| GDF-15 | 1.40 (1.30-1.50) | <2x10^-16^ | 1.38 (1.27-1.51) | 2.8x10^-13^ | 1.25 (1.13-1.39) | 1.7x10^-5^ |
| Hs-CRP | 1.07 (1.00-1.14) | 0.042 | 1.03 (0.97-1.11) | 0.33 | - | - |
| IGFBP-7 | 1.31 (1.22-1.40) | 3.0x10^-14^ | 1.25 (1.16-1.35) | 6.6x10^-9^ | 1.09 (0.99-1.20) | 0.0716 |
| IL-6 | 1.21 (1.13-1.28) | 2.2x10^-9^ | 1.16 (1.09-1.24) | 5.2x10^-6^ | 1.08 (1.01-1.16) | 0.0339 |
| NT-proBNP | 1.29 (1.19-1.40) | 4.2x10^-10^ | 1.21 (1.12-1.32) | 8.8x10^-6^ | 1.06 (0.97-1.17) | 0.21 |
| OPN | 1.24 (1.16-1.32) | 5.4x10^-10^ | 1.19 (1.10-1.29) | 1.8x10^-5^ | - | - |
| hsTropT | 1.25 (1.16-1.33) | 1.6x10^-10^ | 1.19 (1.10-1.28) | 6.5x10^-6^ | - | - |
| *Hazard ratios are standardized per 1-SD increase in biomarker. Multivariable models are adjusted for additional factors controlled for body mass index, current smoker, systolic blood pressure, history of diabetes, prior stroke or TIA, history of heart failure, chronic kidney disease and coronary artery disease. The combined model is adjusted for covariates from the multivariable model plus all significant biomarkers after backward selection based on Akaike Information Criterion (AIC). | | | | | | |

**Supplementary** **Table 13. Biomarkers and risk of clinically relevant non-major bleeding**

|  | **Age and sex adjusted model** | | **Multivariable model** | | **Combined model**  **(backward selection)** | |
| --- | --- | --- | --- | --- | --- | --- |
| **Biomarker** | **HR (95% CI)*** | **P value** | **HR (95% CI)** | **P value** | **HR (95% CI)** | **P value** |
| ANG-2 | 1.24 (1.15-1.35) | 8.8x10^-8^ | 1.20 (1.10-1.30) | 2.7x10^-5^ | - | - |
| eGFR | 0.97 (0.87-1.08) | 0.53 | 0.95 (0.83-1.08) | 0.43 | - | - |
| Cystatin C | 1.15 (1.05-1.25) | 0.0015 | 1.11 (1.01-1.23) | 0.03608 | - | - |
| D-dimer | 1.09 (1.01-1.17) | 0.02 | 1.08 (1.00-1.16) | 0.050 | - | - |
| ALAT | 1.02 (0.94-1.11) | 0.67 | 1.01 (0.92-1.10) | 0.86 | - | - |
| GDF-15 | 1.33 (1.22-1.46) | 5.1x10^-10^ | 1.35 (1.22-1.51) | 3.2x10^-8^ | 1.26 (1.12-1.41) | 0.000135 |
| Hs-CRP | 1.07 (0.99-1.16) | 0.09 | 1.04 (0.96-1.13) | 0.38 | - | - |
| IGFBP-7 | 1.26 (1.15-1.37) | 2.0x10^-7^ | 1.24 (1.12-1.36) | 1.1x10^-5^ | - | - |
| IL-6 | 1.19 (1.10-1.28) | 1.1x10^-5^ | 1.16 (1.07-1.25) | 0.00055 | 1.07 (0.98-1.17) | 0.13 |
| NT-proBNP | 1.31 (1.19-1.45) | 4.6x10^-8^ | 1.26 (1.13-1.40) | 1.7x10^-5^ | 1.15 (1.03-1.29) | 0.013940 |
| OPN | 1.14 (1.04-1.25) | 0.004 | 1.11 (1.00-1.23) | 0.04421 | - | - |
| hsTropT | 1.16 (1.06-1.27) | 0.00099 | 1.11 (1.01-1.23) | 0.0344 | - | - |
| *Hazard ratios are standardized per 1-SD increase in biomarker. Multivariable models are adjusted for additional factors controlled for body mass index, current smoker, systolic blood pressure, history of diabetes, prior stroke or TIA, history of heart failure, chronic kidney disease and coronary artery disease. The combined model is adjusted for covariates from the multivariable model plus all significant biomarkers after backward selection based on Akaike Information Criterion (AIC). | | | | | | |

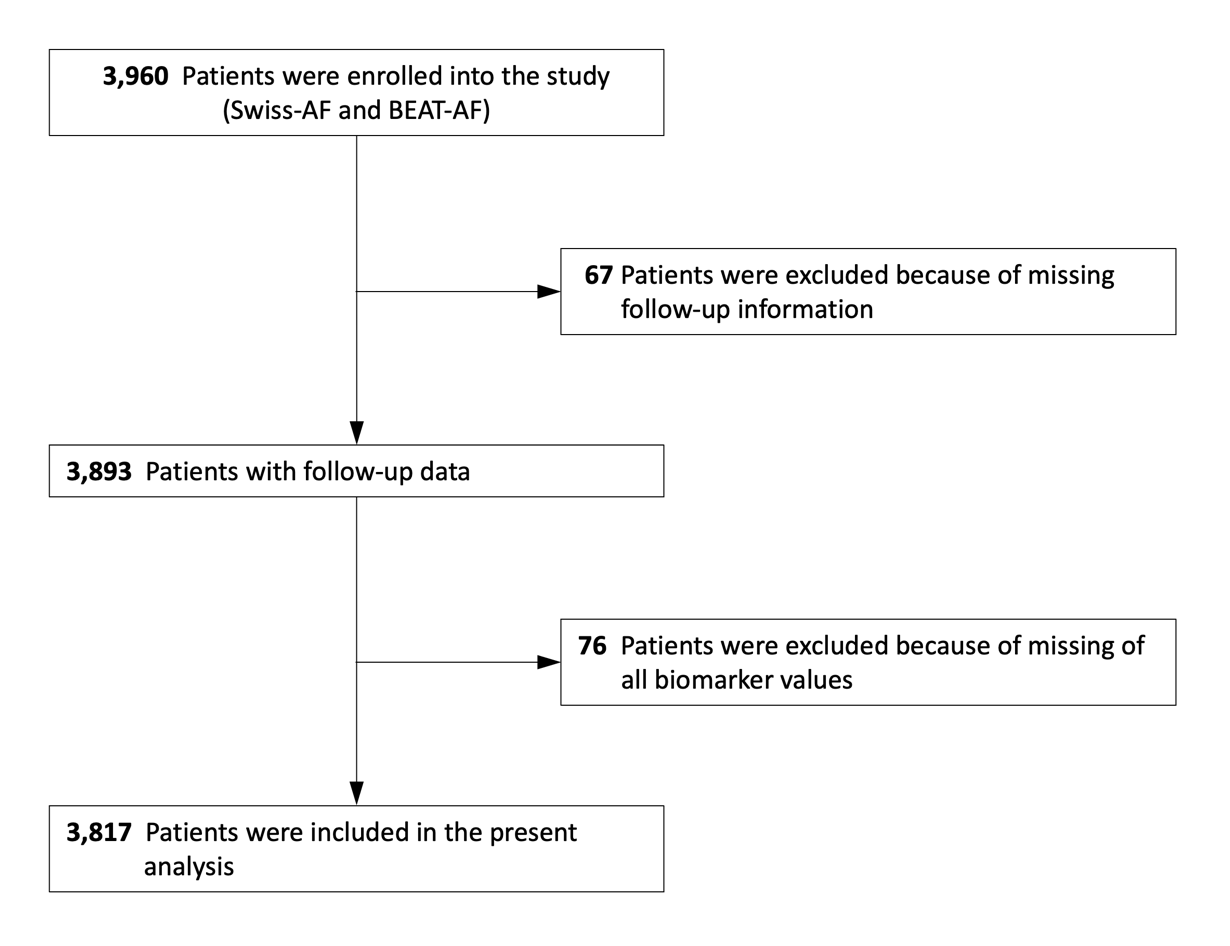
**Supplementary Figure 1. Flow diagram of the study**

**Supplementary Figure 2. Missing pattern and multiple imputation of biomarkers**

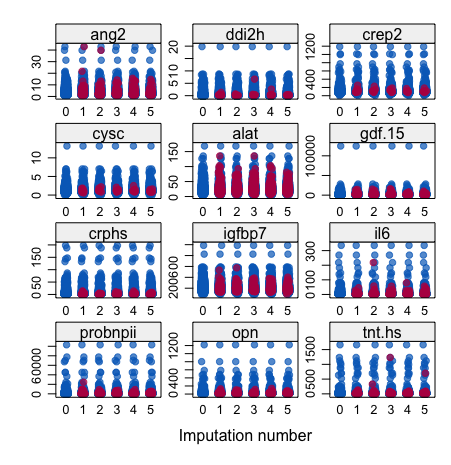

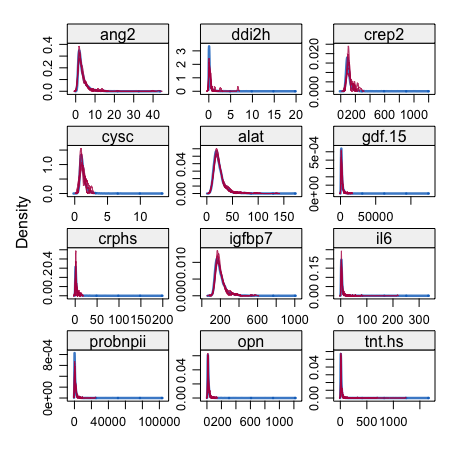

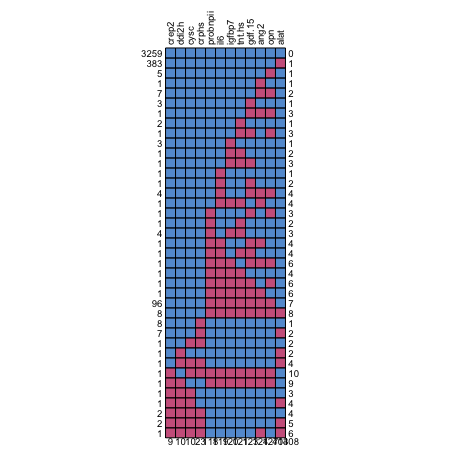
**A**	**B**	**C**

Panel A shows the distribution and extent of missing biomarker values across the dataset. The missing values are highlighted in magenta. Panel B shows the distribution of imputed values of biomarkers from multiple imputations; imputed values are highlighted in magenta. Panel C shows the distribution and variation of data points across different imputations and different biomarkers.

**Supplementary Figure 3. Spearman rank correlations of biomarkers**

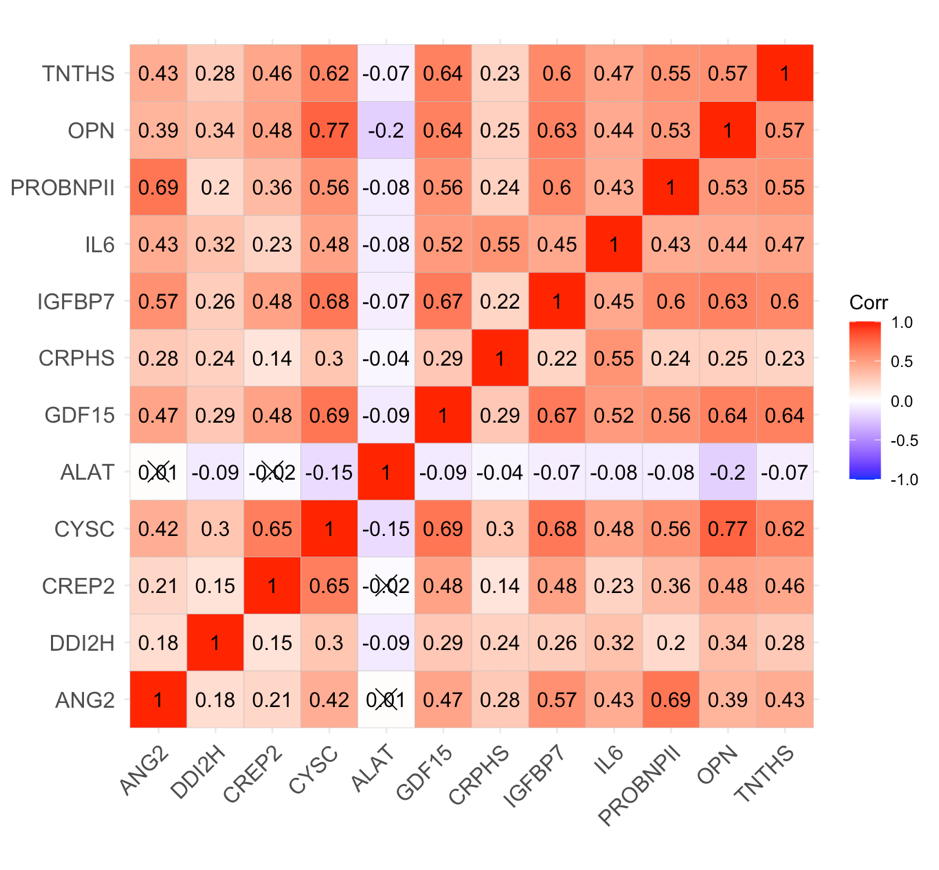

This figure shows the correlations between measured biomarkers. The size and color intensity of each circle correspond to the strength and direction of the correlation.

Abbreviations: ANG2=Angiopoetin-2; DDI2H=D-dimer; CREP2=Creatinine; CYCS=Cystatin C; ALAT=Alanine aminotransferase; GDF-15=Growth differentiation factor‑15; CRPHS= C-reactive protein high sensitive; IGFBP7= Insulin-like growth factor-binding protein-7; IL6= Interleukin-6; PROBNPII=NT-proBNP; OPN=Osteopontin; TNTHS=Troponin T high sensitive.

**Supplementary Figure 4.** Risk of adverse cardiovascular outcomes by biomarkers

This figure shows the risk of adverse cardiovascular outcomes by the 12 measured biomarkers. Risk estimates are from multivariable models adjusted for age, sex, body mass index, current smoker, systolic blood pressure, history of diabetes, prior stroke or TIA, history of heart failure, chronic kidney disease and coronary artery disease. The size and color intensity of each dot correspond to the strength and direction of the association.

**Supplementary Figure 5.** Predictive performance of Cox and machine learning models for outcomes with and without biomarkers
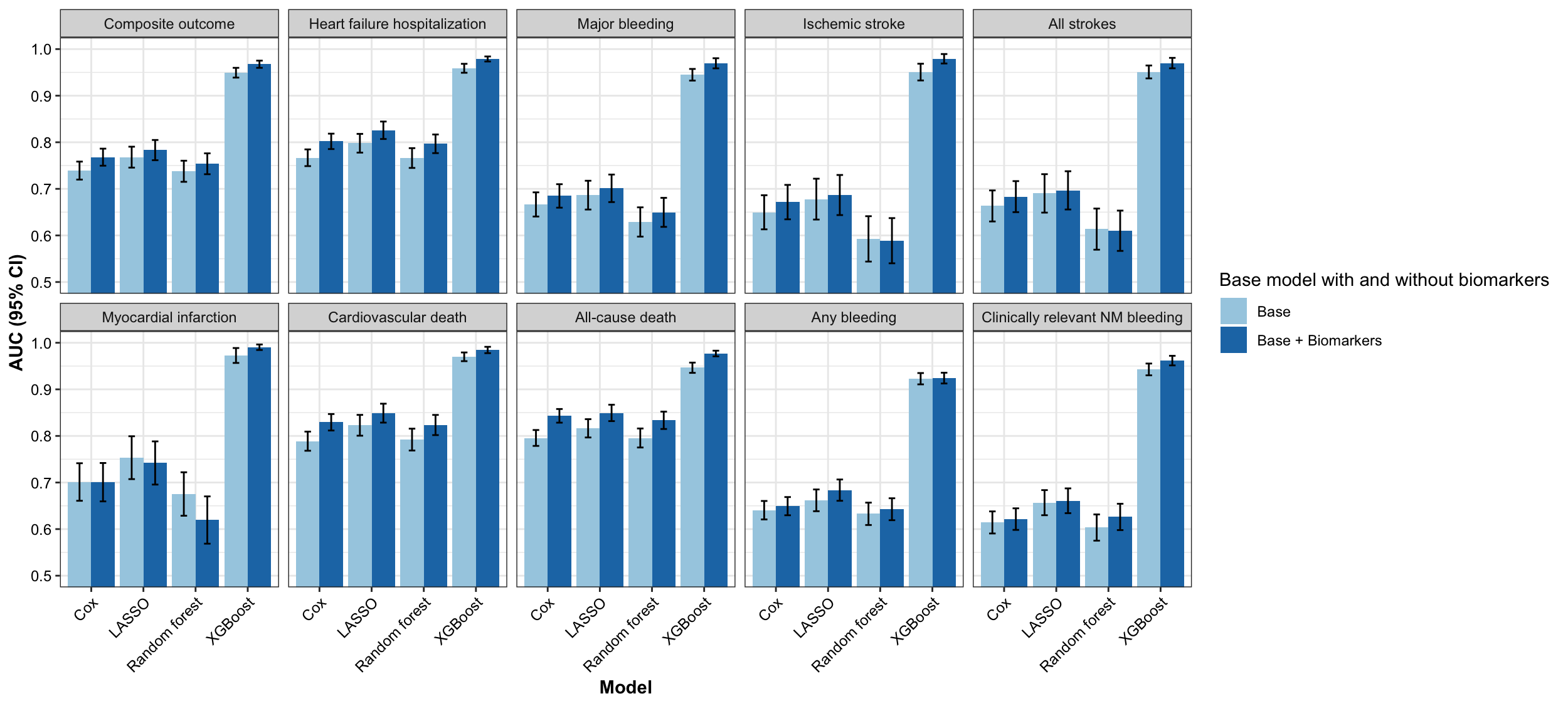

The figure shows the mean AUC with 95% CI for Cox and machine learning models, comparing the performance of base and base + biomarker models for different adverse cardiac outcomes. The combined Cox models include age, sex, body mass index, current smoker, systolic blood pressure, history of diabetes, prior stroke or TIA, history of heart failure, chronic kidney disease, coronary artery disease, and backward-selected biomarkers. The machine learning models include all variables listed in the Supplementary Table 2 and all biomarkers.
